# Response to Polygenic Risk: Results of the MyGeneRank Mobile Application-Based Coronary Artery Disease Study

**DOI:** 10.1101/2021.04.26.21256141

**Authors:** Evan D. Muse, Shang-Fu Chen, Shuchen Liu, Brianna Fernandez, Brian Schrader, Bhuvan Molparia, André Nicolás León, Raymond Lee, Neha Pubbi, Nolan Mejia, Christina Ren, Ahmed El-kalliny, Ernesto Prado Montes de Oca, Hector Aguilar, Arjun Ghoshal, Raquel Dias, Doug Evans, Kai-Yu Chen, Paris Zhang, Nathan E Wineinger, Emily G. Spencer, Eric J. Topol, Ali Torkamani

## Abstract

The degree to which polygenic risk scores (PRS) influence preventive health is the subject of debate, with few prospective studies completed to date. We developed a smartphone application for the prospective and automated generation, communication, and electronic capture of response to a PRS for coronary artery disease (CAD). We evaluated self-reported actions taken in response to personal CAD PRS information, with special interest in the initiation of lipid lowering therapy (NCT03277365). 20% of high genetic risk (n=95) vs 7.9% of low genetic risk individuals (n=101) initiated lipid lowering therapy at follow-up (p-value = 0.002). The initiation of both statin and non-statin lipid lowering therapy was associated with degree of genetic risk – 15.2% (n=92) vs 6.0% (n=100) for statins (p-value = 0.018) and 6.8% (n=118) vs 1.6% (n=123) for non-statins (p-value = 0.022) in high vs low genetic risk, respectively. Overall, degree of genetic risk was associated with use of any lipid lowering therapy at follow-up - 42.4% (n=132) vs 28.5% (n=130) (p-value = 0.009). We also find that CAD PRS information is perceived to be understandable, actionable, and does not induce health anxiety.

## Introduction

Polygenic risk scores (PRS) are promising tools for early risk detection, risk stratification, therapy prioritization, and life-planning (1–3). The utility and effectiveness of polygenic risk estimation in health-decision making has been the recent subject of intense interest and debate (4–7). Early studies exploring the behavioral impact of genetic risk found no significant influence on behavior, though they have generally been performed in a context where specific actionability was limited both due to the broad nature of direct-to-consumer genetic risk panels and the limited risk stratification conveyed by early polygenic risk estimates (8–10). Contemporary, indication-specific and prospective studies on PRS utility are limited.

PRSs for coronary artery disease (CAD) are of special interest given that CAD is a highly heritable condition and the leading cause of preventable death in the developed world (11–14). While there remains some debate regarding the precise utility of CAD PRSs in risk stratification (15–18), high polygenic risk has been independently and repeatedly associated with enhanced benefit from lipid lowering therapy (19–24). This allows a CAD PRS to act as a “risk-enhancer” in any CAD preventive health-decision making scenario, beyond its contribution to risk stratification (14,25,26). Furthermore, only ∼25% of individuals who should use lipid lowering therapies according to clinical guidelines do so, with no bias of use towards individuals with high polygenic risk, thus PRSs could be useful for simply encouraging adherence to clinical guidelines (27).

Beyond multi-disease direct-to-consumer genetic studies (8–10), a few focused clinical studies investigating patient actions in response to CAD PRS have been pursued (28,29). These pilot studies were relatively small (n = 100 – 200) and investigated the clinical profiles of individuals receiving a clinical risk score vs a clinical risk score and PRS (rather than across PRS risk tiers). These studies were also recruited and conducted in clinical settings with direct clinical staff interaction in the communication of risk. Encouragingly, an improvement in clinical profiles was observed for all individuals receiving a PRS, regardless of their degree of genetic risk. A more recent Finnish study, Kardiokompassi, also enhanced with electronic risk visualization, has reported similar preliminary findings supporting a role in risk reduction broadly (30).

Here we describe a prospective, app-based study, *MyGeneRank*, of CAD PRS communication, where participants join through self-referral from the community and provide informed consent electronically. Study participant response to CAD PRS information is self-initiated and self-guided with the support of interactive risk visualizers. Study participant response is gauged through electronic questionnaire. We find that communication of genetic risk through a CAD PRS results in significantly improved alignment of therapeutic risk reduction with the degree of genetic risk in the population. Overall, we find that CAD PRS communication is a promising tool in supporting risk factor optimization for CAD prevention.

## Methods

### Overview

The study protocol “MyGeneRank” was approved by the Scripps IRB Protocol #: IRB-16-6835. The MyGeneRank study was launched in August of 2017 as solely an iOS application. The initial development and basic design is described previously (31). The first version of the study app calculated and returned a 57-SNP CAD PRS based upon the latest CAD GWAS meta-analysis at the time (32). An update was launched in December 2019 expanding the application to Android, adding Spanish language, adding a genetic counseling option, and improving the overall interface. The CAD PRS was also updated to calculate and return a 163-SNP CAD PRS based upon the prior score plus the latest efforts in identifying potential causal alleles (33) and weights from the latest CAD GWAS meta-analysis (34). The calculation of this PRS is previously described in detail (35). Screenshots of the current app interface can be viewed at the study website: mygenerank.scripps.edu. Most scores did not change substantially after the update, details of which can be found here: mygenerank.scripps.edu/blog/post/mgr-new-update.

### Study Tasks

Adults (≥18 years old) with existing genetic data from 23andMe were eligible to participate in the MyGeneRank study. Participants download the study application and are presented with an eligibility screen and series of screens summarizing the consent, a complete consent form, HIPAA authorization, and participant bill of rights. PDF copies of these documents are available to study participants within the app, and participants may withdraw from the study at any time. Study participants are asked to link their 23andMe genetic data via API, optionally provide access to their mobile health data via Apple Health and Google Fit, and answer three survey questionnaires. These questionnaires include: (1) a 28-question initial demographic and CAD health and history survey answered prior to the return of results, (2) a 14-question psychosocial survey delivered immediately after the return of results aimed at gauging study participant intentions and feelings in response to their CAD PRS, and (3) a follow-up CAD health survey with questions overlapping with the initial healthy and history survey. The follow-up survey is made available to study participants in the study app at 6-months post-results but could be answered at any time after 6-months post-results. Follow-up responses were also obtained via email-based electronic survey disseminated in November 2020 to January 2021. 3 questions were added to the baseline and follow-up survey, and 4 questions to the psychosocial survey with the December 2019 update.

### Enrollment and Responses

A total of 3,800 valid participants, after duplicate removal, connected their existing 23andMe genetic data with the MyGeneRank system and at least partially answered the initial survey questionnaire. Approximately, 80% of study participants joined the study prior to the December 2019 update. Of the individuals enrolling prior to the score update, 2.5% responded to the psychosocial survey and 67% responded to the follow-up CAD health survey after receiving their updated score. 253 (6.7%) of participants responded proactively to the app-based follow-up survey. 459 (12%) of participants responded to an email-based follow-up survey after re-contact. Missing responses lead to small differences in the number of study respondents answering each individual question. Herein, we refer to “study participants” as any enrolled individual. “Study respondent” refers to those enrolled individuals whose response includes the final follow-up survey.

### Risk Communication

Study participants view their CAD risk in two interfaces: (1) an interface integrating the CAD PRS with the 10-year ASCVD Pooled Cohort Equations derived risk, and (2) a percentile rank view where the percentile rank is calculated relative to simulated individuals of similar genetic ancestry (31). Study participants are categorized as low (<20^th^ percentile), average (20^th^ to 80^th^ percentile), or high (>80^th^ percentile) genetic risk. Any additional participant actions, including adoption of genetic counseling services, sharing of results with a physician, or any other actions taken outside of score receipt and response to study surveys were unprompted and self-initiated by study participants.

### Comparisons

For comparisons involving survey responses at baseline vs follow-up, the score the participant received at baseline is used for categorization. For comparisons involving survey responses at a single timepoint, the score (original vs updated) available to the participant at the time of their response was used. Comparisons are made with a standard one-tailed two-proportion z-test for high vs low genetic risk individuals. Likert score responses were made with a two-tailed Mann Whitney U test for high vs low genetic risk individuals. Likert scores were coded as 1: strongly disagree, 2: somewhat disagree, 3: neutral, 4: somewhat agree, 5: strongly agree.

## Results

### Study Participant Baseline Characteristics

Participant characteristics at baseline are presented in **Table 1**. For the subset of individuals providing complete information, we determined their clinical risk using the Pooled Cohort Equation (26). The study population is largely comprised of clinically low-risk (< 7.5% 10-year ASCVD risk based on PCE) individuals of European ancestry, with 73% of study participants and 63% of study respondents belonging to the clinical low-risk category at enrollment. Encouragingly, study respondents were significantly enriched with individuals in the intermediate clinical risk category (13% of study participants vs 20% of study respondents), which corresponds to the clinical risk category where CAD PRS information could be most useful as a risk-enhancing factor mediating the initiation or titration of lipid lowering therapy. This enrichment in intermediate clinical risk was associated with an increased response from older individuals as opposed to enriching for negative modifiable CAD risk factor characteristics (**Table 1**). The other major difference in participants vs respondents was the use of lipid lowering therapy at baseline: 16% of study participants vs 27% of study respondents report taking a statin at baseline. Similarly, 5.7% vs 9.8% of study participants vs respondents report the use of other (non-statin) lipid lowering therapies at baseline. Thus, study respondents remain largely low risk but are enriched with intermediate risk individuals and those taking lipid lowering medications.

**Table 1.** Study Participant and Respondent Characteristics.

|  | Study Participants<br>(n = 3,484) | Study Respondents<br>(n = 712) |
| --- | --- | --- |
| **Gender** (% female) | 49% | 42% |
| **Age** |  |  |
| 18 - 29 | 17.4% | 8.3% |
| 30 - 39 | 31.2% | 23.6% |
| 40 - 49 | 22.1% | 21.4% |
| 50 - 59 | 16.5% | 22.2% |
| 60 - 69 | 9.6% | 17.6% |
| 70+ | 3.2% | 7.4% |
| **Ancestry** |  |  |
| African American | 1.9% | 1.3% |
| American and Alaskan Native | 1.6% | 0.9% |
| Asian | 5.5% | 3.4% |
| Pacific Islander | 0.6% | 0.3% |
| White | 92.9% | 95.2% |
| Decline | 1.5% | 1.5% |
| **Ethnicity** (% Hispanic or latino) | 6.8% | 5.6% |
| **Body Mass Index** |  |  |
| < 20 | 9.1% | 8.5% |
| 20 – 25 | 21.9% | 21.4% |
| 25 – 30 | 38.4% | 39.9% |
| 30 – 35 | 24.0% | 25.6% |
| ≥ 35 | 6.6% | 6.0% |
| **CAD Risk Factors (% self-reported)** |  |  |
| high cholesterol | 13% | 15% |
| high blood pressure | 4.6% | 4.9% |
| smoking | 6.4% | 2.6% |
| diabetes | 2.7% | 3.4% |
| active lifestyle | 72% | 76.5% |
| healthy diet | 62% | 63% |
| statin medication | 16% | 27% |
| other lipid lowering | 5.7% | 9.8% |
| **10-year ASCVD Risk** | (n = 990) | (n = 282) |
| < 5% | 73% | 63% |
| 5 – 7.5% | 10% | 11% |
| 7.5% - 20% | 13% | 20% |
| > 20% | 4% | 6% |

### Study Participant Reactions

Of the 3,800 study participants, 1,053 (28%) provided their immediate reaction to learning their CAD PRS by expressing their degree of (dis)agreement with 12 statements. The statements where reactions differed significantly by CAD PRS category are presented in **Table 2**. All statements with large, standardized effects are related to self-perceptions of risk: i.e. “My chances of developing Coronary Artery Disease are high” and “My genetics make it more likely that I will get coronary artery disease” – where low PRS individuals expressed mild disagreement, average PRS individuals were neutral, and high PRS individuals expressed agreement with these statements. A small effect was also observed for: “I worry a lot about developing Coronary Artery Disease”, where low and average PRS individuals expressed mild disagreement, and high PRS individuals were neutral in response to this statement. Reactions to other statements are provided in **Supplemental Table 1**. Overall, study participants agreed uniformly that they understood their risk, that they are able to reduce their risk, and that CAD is a serious condition.

**Table 2.** Study Participant Reactions.

| Statement | n | Low Genetic Risk | Average Genetic Risk | High Genetic Risk | Standardized Effect Size | P-value |
| --- | --- | --- | --- | --- | --- | --- |
| **My chances of developing Coronary Artery Disease are high.** | low=69, avg=165, high=73 | 2.39 ± 0.14 | 3.13 ± 0.09 | 4.18 ± 0.13 | 0.63 | 8.8E-14 |
| **My genetics make it more likely that I will get Coronary Artery Disease.** | low=69, avg=165, high=73 | 2.48 ± 0.16 | 3.33 ± 0.09 | 4.23 ± 0.12 | 0.61 | 3.08E-13 |
| **My chances of getting Coronary Artery Disease are high.** | low=170, avg=406, high=152 | 2.25 ± 0.09 | 2.96 ± 0.06 | 3.86 ± 0.09 | 0.58 | <1e-16 |
| **I am at risk of getting Coronary Artery Disease.** | low=170, avg=406, high=152 | 2.84 ± 0.1 | 3.53 ± 0.06 | 4.15 ± 0.08 | 0.49 | 1.11E-16 |
| **I worry a lot about developing Coronary Artery Disease.** | low=239, avg=571, high=224 | 2.44 ± 0.08 | 2.68 ± 0.05 | 3.09 ± 0.08 | 0.25 | 7.38E-08 |

Psychosocial responses showing significant difference across CAD PRS tiers. Average Likert scores plus standard error of the mean are displayed. Significance determined by Mann Whitney U test.

### Study Participant Intentions

Similarly, study participants were surveyed for their intended actions after receiving their CAD PRS. Specifically, participants were asked (1) “Now that you know your coronary artery disease genetic risk score, do you intend to make any changes to your use of statins?”, and (2) “Now that you know your coronary artery disease genetic risk score, do you intend to meet with a physician to discuss these results?” Study participants reported intentions to meet with their physician (2-fold difference in high vs low genetic risk) and begin the use of statins (4-fold difference in high vs low genetic risk) in strong association with their degree of genetic risk (**Table 3**)

**Table 3.** Study Participant Intentions.

| **Statement** | **Low**<br>**Genetic**<br>**Risk**<br>**(n = 232)** | **Average**<br>**Genetic**<br>**Risk**<br>**(n = 581)** | **High**<br>**Genetic**<br>**Risk**<br>**(n = 240)** | **P-value** |
| --- | --- | --- | --- | --- |
| **Now that you know your coronary artery disease genetic risk score, do you intend to make any changes to your use of statins?**<br>**(% begin or increase use of statins)** | 5% | 10.5% | 19.0% | < .00001 |
| **Now that you know your coronary artery disease genetic risk score, do you intend to meet with a physician to discuss these results? (% yes)** | 33.3% | 40.8% | 60.3% | < .00001 |

Study participant intentions after receipt of their CAD PRS. Significance determined by two proportion z-test of high vs low risk individuals.

### Study Respondent Actions

At 6-months follow-up or later, 253 study respondents proactively answered this survey within the study app while 459 study respondents answered the email survey. Follow-up times were 1-year on average for app-based respondents and 1.5 years on average for email-based respondents. We gauged the initiation and/or intensification of lipid lowering in two ways: 1) by comparing responses relating to lipid lowering therapy use at baseline and follow-up, and 2) by asking about attribution of changes in lipid lowering to receipt of the PRS at follow-up.

For our primary analysis, we compared baseline and follow-up responses to two questions: (1) “Are you currently taking or have you previously taken a statin?” and (2) “Are you currently taking or have you previously taken any medication, other than a statin, used to treat high cholesterol?” The comparison of baseline vs follow-up responses is presented in **Table 4**. Study respondents of high genetic risk and not taking a statin at baseline, initiated statin therapy at ∼2.5-fold the rate of low genetic risk study respondents (15.2% vs 6% statin initiation rate, p-value = 0.018). Similarly, study respondents of high genetic risk and not taking non-statin lipid lowering therapy at baseline, initiated non-statin lipid lowering therapy at ∼3-fold the rate of low genetic risk individuals (6.8% vs 1.6% non-statin initiation rate, p-value 0.022). Discontinuation rates were low and not associated with degree of genetic risk (**Table 4**).

**Table 4.** Initiation and Discontinuation of Lipid Lowering Therapy.

|  |  |  | Low Genetic Risk<br>(n = 132) |  | Average Genetic Risk<br>(n = 387) |  | High Genetic Risk<br>(n = 130) |  | Initiation Rate<br>(p-value) |
| --- | --- | --- | --- | --- | --- | --- | --- | --- | --- |
|  |  |  | Follow-up |  | Follow-up |  | Follow-up |  | low, avg, high |
|  |  |  | No | Yes | No | Yes | No | Yes |  |
| Statins | Baseline | No | 72.4% | **4.6%** | 69% | 4.4% | 59% | **10.6%** | 6%, 6%, 15.2%<br>(0.018) |
|  |  | Yes | 0% | 23% | 2.1% | 24.5% | 0.8% | 29.5% |  |
| Other Lipid Lowering | Baseline | No | 93.1% | **1.5%** | 84.5% | 3.6% | 83.3% | **6.1%** | 1.6%, 4.1%, 6.8%<br>(0.022) |
|  |  | Yes | 3.8% | 1.5% | 5.9% | 5.9% | 3.8% | 6.8% |  |

Study respondent responses to use of statin and non-statin lipid lowering at baseline and follow-up. Values of most interest are bolded and italicized. Significance determined by two proportion z-test of high vs low risk individuals.

We also asked directly about lipid lowering actions attributed to receipt of the CAD PRS through the following three questions: (1) “Did you meet with a physician to discuss your coronary artery disease risk score?,” (2) “After receiving your coronary artery disease genetic risk score, did you make any changes to your use of statins?,” and (3) “After receiving your coronary artery disease genetic risk score, did you make any changes to your use medications, other than statins, used to treat high cholesterol?” In response to this question, a clear difference in response to the PRS by level of engagement was apparent (**Table 5**). Proactive app-based responses displayed a significant relationship between attribution of changes in lipid lowering therapy to receiving a PRS, while email-based responses did not maintain this relationship. Study respondents reported that they met to discuss their results with their physician regardless of degree of risk, with a non-significant trend in association with degree of genetic risk.

**Table 5.**
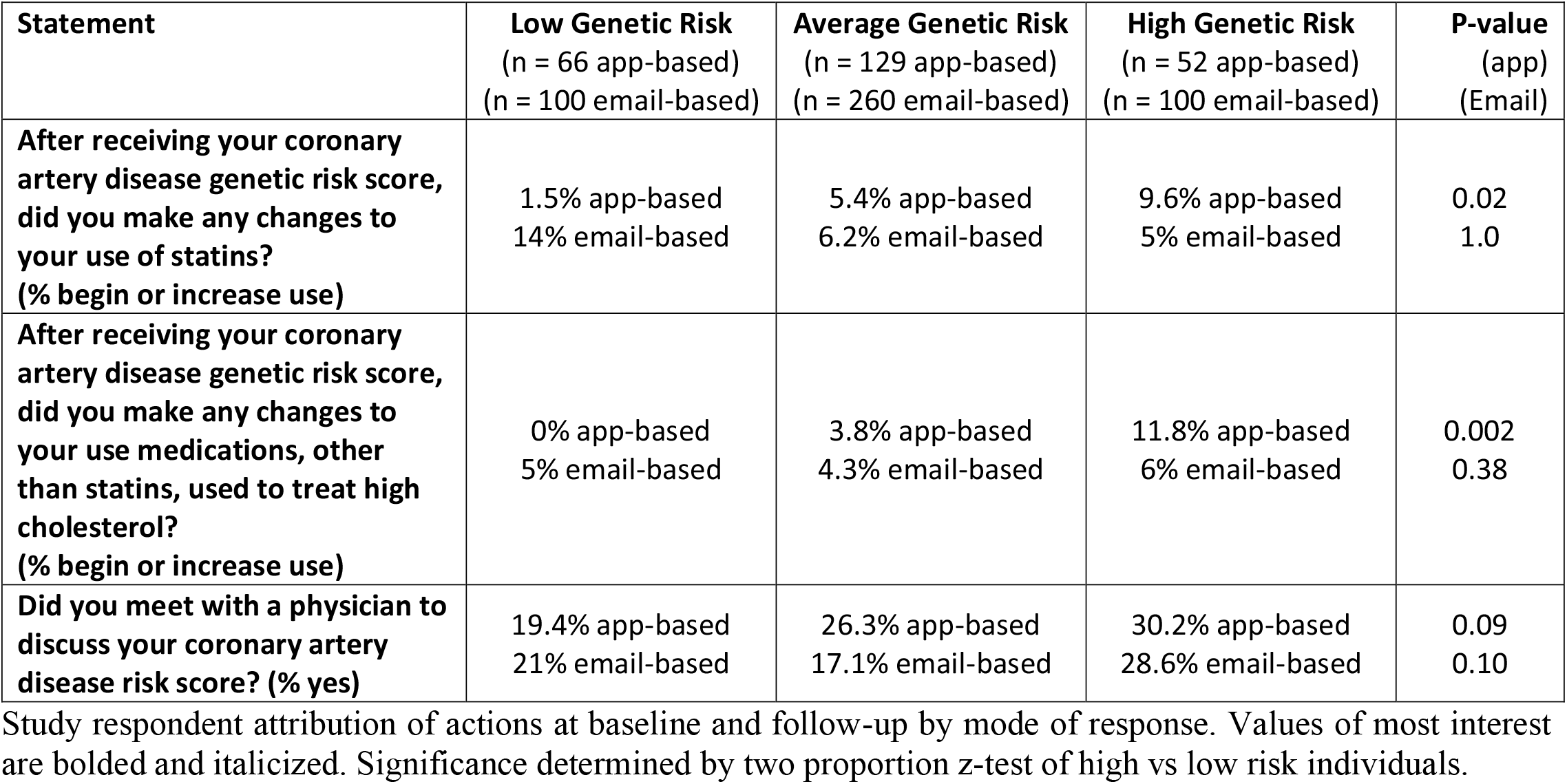
Attribution of Lipid Lowering Therapy Changes.

| **Statement** | **Low Genetic Risk**<br>(n = 66 app-based)<br>(n = 100 email-based) | **Average Genetic Risk**<br>(n = 129 app-based)<br>(n = 260 email-based) | **High Genetic Risk**<br>(n = 52 app-based)<br>(n = 100 email-based) | **P-value**<br>(app)<br>(Email) |
| --- | --- | --- | --- | --- |
| **After receiving your coronary artery disease genetic risk score, did you make any changes to your use of statins? (% begin or increase use)** | 1.5% app-based<br>14% email-based | 5.4% app-based<br>6.2% email-based | 9.6% app-based<br>5% email-based | 0.02<br>1.0 |
| **After receiving your coronary artery disease genetic risk score, did you make any changes to your use medications, other than statins, used to treat high cholesterol? (% begin or increase use)** | 0% app-based<br>5% email-based | 3.8% app-based<br>4.3% email-based | 11.8% app-based<br>6% email-based | 0.002<br>0.38 |
| **Did you meet with a physician to discuss your coronary artery disease risk score? (% yes)** | 19.4% app-based<br>21% email-based | 26.3% app-based<br>17.1% email-based | 30.2% app-based<br>28.6% email-based | 0.09<br>0.10 |

Study respondent attribution of actions at baseline and follow-up by mode of response. Values of most interest are bolded and italicized. Significance determined by two proportion z-test of high vs low risk individuals.

### Overall Use of Lipid Lowering Therapies

Finally, we compare the rate of statin, non-statin, and combined lipid lowering therapies at baseline and follow-up across study respondents (**Table 6**). While we observe a baseline statin usage rate of 27% overall, which is consistent with prior reports (27), our results conflict with prior reports in that we observe a weak association of lipid lowering with degree of genetic risk at baseline (**Table 6**). This association achieves statistical significance only for combined use of lipid lowering therapies. At follow-up, a stronger and significant association of statin, non-statin, and combined lipid lowering therapy use with degree of genetic risk is observed. Study respondents with high genetic risk were ∼1.4-fold more likely to report use of a statin at follow-up and ∼4-fold more likely to report use of a non-statin lipid lowering medication at follow-up, resulting in a ∼1.5X difference in the rate of combined lipid lowering therapy use in high vs low genetic risk individuals (**Table 6**). When considering only study respondents not engaging in any lipid lowering therapy at baseline, we observe a strong and significant association of lipid lowering therapy initiation with degree of genetic risk - 7.9% (n = 101) vs 9.9% (n = 284) vs 20% (n = 95) of study respondents initiate any lipid lowering therapy in low, average, and high genetic risk individuals respectively (p-value = 0.002, two proportion z-test). Thus, the overall rate of initiation of lipid lowering therapy was >2-fold in study respondents receiving a high vs low CAD PRS.

**Table 6.**

|  | **Low Genetic Risk**<br>(n = 132) |  | **Average Genetic Risk**<br>(n = 387) |  | **High Genetic Risk**<br>(n = 130) |  | **P-value** |
| --- | --- | --- | --- | --- | --- | --- | --- |
|  | Baseline | Follow-up | Baseline | Follow-up | Baseline | Follow-up | (Baseline)<br>(Follow-up) |
| **Statins** | 23.1% | **27.7%** | 26.6% | 28.9% | 30.3% | **40.2%** | 0.09<br>**0.017** |
| **Non-statin** | 5.4% | **3.1%** | 11.9% | 9.6% | 10.6% | **12.9%** | 0.06<br>**0.002** |
| **Combined Lipid Lowering** | 23.1% | **28.5%** | 29.2% | 31.5% | 33.3% | **42.4%** | 0.03<br>**0.009** |

Study respondent use of statin and non-statin lipid lowering at baseline and follow-up. Values of most interest are bolded and italicized.

## Discussion

Here we report the results of a real-world, fully-digitized, and participant-centric approach to communication of CAD genetic risk via a PRS combined with clinical risk evaluation. We find that communication of a CAD PRS is effective at improving the alignment of risk reducing interventions with degree of genetic risk. The rate of lipid lowering therapy initiation (20%) in high genetic risk individuals is remarkable given the overall low clinical risk profile of study participants, at least according to the Pooled Cohort Equation.

Our pioneering digital approach to PRS communication is likely a major contributor to the beneficial impact on risk-reducing behaviors we observe for MyGeneRank participants relative to prior studies (8–10). One major advantage is the dynamic and interactive nature of risk communication in the mobile app interface. Rather than making overt suggestions to participants, the dynamic risk reducer interface presents participants with a self-guided, interactive system to contextualize their genetic risk, allowing for a self-efficacious approach to health decision making and risk factor optimization. The MyGeneRank app also acted as an always available digital reference, allowing participants to re-evaluate their risk at the time a health decision is being made. In fact, we designed the backend capabilities so that an individual with existing genetic data can request and receive their CAD PRS in the timespan of a medical office visit. We suggest that self-guided risk contextualization is a key component of PRS utility.

With this contextualization, we find that study participants understand their risk and report self-efficacy in reducing their risk, while minimally impacting health anxiety or worry. While the lack of health anxiety overall could be attributed to a central tendency bias in the Likert score responses, we do not observe a central tendency bias for any other statement, suggesting a true lack of worry or health anxiety. This finding may be specific to CAD, as prior work suggests an overall underappreciation of the risks associated with CAD vs other conditions like cancer (36). Regardless, these results suggest that for CAD, genetic risk can be communicated effectively without adverse psychological effects while aligning risk reducing behaviors with individual genetic risk.

While we observed a strong association of both use and initiation of lipid lowering therapy with genetic risk, there are indications that this effect can be further amplified by deeper engagement with participants and/or with treating physicians as well as genetic counselors. We initially launched the MyGeneRank app with a basic interface and PRS. Additional capabilities such as the link to genetic counseling services, an advanced risk reduction interface with integrated genetic and clinical ASCVD risk, and an updated score were launched after the December 2019 update. Individual level changes in the score (35), as well as access to different capabilities may have influenced the consistency of our results, however these considerations would be expected to diminish the association of the CAD PRS with risk reducing actions. Differences in attribution of response from app-based vs email-based responses suggest the influence of CAD PRS communication may diminish overtime if not reinforced, though the ultimate enhanced rate of lipid lowering in high genetic risk individuals at follow-up does not bear out this concern. Another limitation includes the study population, which likely consists of early adopters who may be more engaged in preventive health in comparison to a more general population. Additionally, use of the mobile app was not restricted based on existing atherosclerotic heart disease and the proportion of participants who were using this to inform their strategies of primary versus advanced secondary prevention is not known. Finally, ∼1-year of this study overlaps with the ongoing SARS-CoV-2 pandemic, which has reportedly decreased the use of primary care interventions overall, including cholesterol testing (37).

Overall, we find that the communication of a CAD PRS is highly effective at aligning risk-reducing behaviors with degree of risk. A clear and strong association of the communication of high genetic risk for CAD with initiation of new lipid lowering therapy is observed. Further objective outcomes, including those derived from EHR, are required to confirm this finding and to dissect the role of CAD PRS communication in intensification of lipid lowering therapy in individuals taking sub-optimal statin dose or other lipid lowering medications. The use of CAD PRS should be strongly considered in clinical-decision making scenarios as a risk enhancer where a high genetic risk score could reinforce or enhance guideline-based recommendations, especially in the face of poor adherence to these guidelines overall. Longer term objective studies are required to determine whether communication of a CAD PRS can extend its influence from initiation to intensification, adherence, and persistence of lipid lowering therapy, as well as improved hard outcomes. Finally, we suggest that PRS utility studies should be executed in indication-specific contexts to accurately gauge their influence on early prevention.

## Data Availability

Raw study data will be made available upon final publication at the publishing journals website via supplementary materials.

## List of Abbreviations

CAD: Coronary artery disease
GWAS: Genome-wide association study
PRS: Polygenic risk score
SNP: Single-nucleotide polymorphism
avg: average

## Declarations

### Ethics approval and consent to participate

The study protocol “MyGeneRank” was approved by the Scripps IRB Protocol #: IRB-16-6835. Informed consent was collected electronically in the study app. This research conforms with the principles of the Helsinki Declaration.

### Consent for publication

Individuals were consented for publication at enrollment.

### Availability of data and materials

The raw study data supporting all tables is available as Supplemental Table 1. Individual-level genetic data is not available for sharing and was not generated with NIH funding.

### Code availability

Code used to generate polygenic risk scores is available at via the Torkamani lab github repository: https://github.com/TorkamaniLab

### Authors’ contributions

EDM, EGS, and EJT conceived and designed the study and contributed to manuscript writing. SFC, SL, BF, BS, BM, ANL, RL, NP, NM, CR, AE, EPMO, HA, AG, RD, DE, KC, PZ and NEW collected data and contributed data tools (developed components of the app). AT conceived and designed the study, analyzed and interpreted study data, and contributed to manuscript writing. All authors read and approved the final manuscript.

## Acknowledgements

We would like to thank J.C. Ducom, Lisa Dong, and the Scripps High Performance Computing service for their support. We would also like to thank DNAVisit for their genetic counseling support and 23andMe for maintaining their API access.

## Supplementary Information

**Additional file 1(**.**xlsx): Supplementary Tables S1-S3**.

Raw Study Data (Table S1). Data Definition Table (Table S2). Psychosocial Response (Table S3).

